# Sub-national estimates of vitamin A supplementation coverage in children: a geospatial analysis of 45 low and middle-income countries

**DOI:** 10.1101/2023.05.09.23289711

**Authors:** Jacqueline Seufert, Nandita Krishnan, Gary L. Darmstadt, Grace Wang, Till Bärnighausen, Pascal Geldsetzer

## Abstract

Vitamin A supplementation (VAS) can protect children from the adverse health consequences of vitamin A deficiency. To inform the geographically precise targeting of VAS programs and provide a benchmark for monitoring progress in reducing geographic disparities in coverage over time, we created high resolution maps (5km x 5km) of the proportion of preschool-age children (6-59 months) covered by VAS in 45 UNICEF designated VAS priority countries using data from the Demographic and Health Surveys program. In addition to prevalence, we estimated absolute VAS coverage and exceedance probabilities using thresholds of 0.5 and 0.7. We found that most countries had coverage levels below 70%. Coverage varied substantially between and within countries. Inter-national variations were most notable in Latin America and the Caribbean, as well as Africa, whereas intra-national variations were greatest in some south Asian and west and central African countries. These maps, especially when used along with high-resolution data on indicators of VAS need, could help VAS programs improve equity.

## 1 Introduction

Vitamin A is an essential nutrient that plays a crucial role in growth and development, immune function, and vision (Tanumihardjo et al 2016). As the human body does not produce vitamin A, it must be obtained through dietary sources. Inadequate dietary intake of vitamin A results in vitamin A deficiency (VAD), which has particularly adverse effects on the health of infants and young children (UNICEF 2018, West Jr 2003). Approximately 29% of preschool-age children (i.e., between the ages of 6-59 months) in low and middle-income countries (LMICs) are estimated to have VAD, defined as serum retinol concentrations *<* 0.70 *μmol/L*; the prevalence of VAD is especially high in sub-Saharan Africa (48%) and South Asia (44%) (Stevens et al 2015).

VAD is thought to be the leading preventable cause of childhood blindness globally (UNICEF 2021). Additionally, VAD increases susceptibility to infectious diseases, such as diarrhea and measles (UNICEF 2018, West Jr 2003). Diarrhea is among the top five leading causes of death in children under 5 years (∼ 450,000 deaths) (Perin et al 2022, Troeger et al 2018) and roughly 16.5% of these deaths are estimated to be attributable to VAD (Stevens et al 2015). Measles is also a leading cause of child deaths (∼ 180,000 deaths) (Perin et al 2022) and VAD is estimated to account for approximately 11% of measles deaths among children in LMICs (Stevens et al 2015). Notably, the burden of diarrhea and measles among children is greatest in the regions with the highest prevalence of VAD (i.e., sub-Saharan Africa and South Asia) (Troeger et al 2018, Perin et al 2022). Thus, addressing VAD is thought to have a marked effect on child health and survival.

Vitamin A supplementation (VAS) is a low-cost population level intervention that protects against the adverse effects of VAD (UNICEF 2018). VAS has been shown to substantially reduce morbidity and mortality in pre-school age children (Imdad et al 2022) and the World Health Organization (WHO) recommends biannual high-dose supplementation for pre-school age children in settings where VAD is a public health problem (WHO 2011). The United Nations Children’s Fund (UNICEF) identified 82 priority countries for national VAS programming in 2000 (UNICEF 2021), based on their VAD prevalence and under-5 mortality rate (U5MR), a surrogate indicator of VAD (McLean et al 2020, Schultink 2002). While universal coverage is the goal, a coverage rate ≥ 70% is considered the minimum threshold at which reductions in child mortality can be observed (UNICEF 2007).

VAS coverage in priority countries increased dramatically, from 27% in 2000 to an all-time high of 78% in 2009; however, global coverage rates have since declined (UNICEF 2018). Concerningly, declines in coverage have been greatest in countries with the highest child mortality rates, indicating that children with the greatest need for VAS are not receiving it (UNICEF 2018). VAS coverage may also be uneven within countries and children living in the most vulnerable communities tend to be disproportionately impacted, further exacerbating inequalities. For instance, children living in rural (vs urban) areas and from poorer families are less likely to receive VAS (Berde et al 2019, Haselow et al 2022, Zegeye et al 2022).

Tracking VAS coverage at a geographically granular level could inform interventions by pinpointing which communities have particularly low coverage, thus contributing to more efficient use of scarce resources. While prior studies have estimated sub-national VAS coverage in select priority countries (Bora 2022, Gilano et al 2021, Zegeye et al 2022), to our knowledge, no comprehensive analysis of VAS coverage data across priority countries has been undertaken. The present study used data from the USAID Demographic and Health Surveys (DHS) program to examine the state of VAS coverage on a granular level in priority countries with available data, an important step towards quantifying and addressing national and sub-national disparities. Using advanced geospatial modelling techniques, we created high-resolution maps (5x5 *km*^2^ pixel) characterizing VAS coverage in 45 priority countries.

## 2 Methods

### 2.1 Surveys and VAS coverage data

For this analysis, we used country-specific surveys from the DHS (ICF 2006-2018). DHS are nationally representative surveys that collect data on a variety of health and nutrition indicators from different countries using standardized methodology; thus, data are comparable across countries. In general, DHS sampling is based on a stratified two-stage cluster design; enumeration areas (EA) obtained from census files comprise the first stage and within each selected EA, sample households make up the second stage. We included countries that were designated VAS priority countries between 2000-2017 by UNICEF. For each country, we used the most recent survey that did not date back further than 2005. A summary of survey characteristics by country can be found in the appendix (see Table 1).

**Table 1:** Survey characteristics by country

| Country | Survey Year | $\phi$ | Number of women | Number of births in the last five years | Household response rate | Women's response rate |
| --- | --- | --- | --- | --- | --- | --- |
| Bangladesh | 2014 | 61.6 | 6,855 | 7,886 | 98.5 | 97.9 |
| Benin | 2017-18 | 52.7 | 8,994 | 13,589 | 99.0 | 98.1 |
| Bolivia | 2008 | 17.7 | 6,429 | 8,605 | 98.8 | 95.9 |
| Burkina Faso | 2010 | 65.3 | 10,364 | 15,045 | 99.2 | 98.4 |
| Burundi | 2016-17 | 66.9 | 8,660 | 13,192 | 99.7 | 98.8 |
| Cambodia | 2014 | 64.0 | 5,901 | 7,165 | 99.3 | 97.6 |
| Cameroon | 2018 | 50.7 | 6,463 | 9,733 | 99.4 | 98.2 |
| Chad | 2014-15 | 41.3 | 11,104 | 18,623 | 98.9 | 96.1 |
| Comoros | 2012 | 48.6 | 2,016 | 3,150 | 96.3 | 93.2 |
| Democratic Republic of Congo | 2013-14 | 64.7 | 11,293 | 18,716 | 99.9 | 98.6 |
| Côte d'Ivoire | 2011-12 | 63.7 | 5,431 | 7,776 | 98.1 | 92.7 |
| Egypt | 2014 | 18.2 | 11,495 | 15,848 | 98.4 | 99.4 |
| Eswatini | 2006-07 | 78.9 | 2,136 | 2,812 | 95.2 | 94.1 |
| Ethiopia | 2016 | 49.6 | 7,193 | 10,641 | 97.6 | 94.6 |
| Gabon | 2012 | 59.9 | 4,143 | 6,067 | 99.3 | 98.2 |
| Ghana | 2014 | 62.2 | 4,294 | 5,884 | 98.5 | 97.3 |
| Guatemala | 2014-15 | 47.4 | 9,542 | 12,440 | 98.7 | 96.8 |
| Guinea | 2018 | 41.1 | 5,530 | 7,951 | 99.2 | 99.0 |
| Haiti | 2016-17 | 31.7 | 5,005 | 6,530 | 99.7 | 98.9 |
| Honduras | 2011-12 | 66.5 | 8,715 | 10,888 | 98.3 | 93.2 |
| India | 2015-16 | 56.4 | 190,898 | 259,627 | 97.6 | 96.7 |
| Kenya | 2014 | 69.3 | 14,949 | 20,964 | 99.0 | 96.6 |
| Kyrgyz Republic | 2012 | 44.9 | 3,148 | 4,363 | 99.5 | 99.1 |
| Lesotho | 2014 | 55.7 | 2,596 | 3,138 | 98.5 | 97.1 |
| Liberia | 2013 | 55.5 | 5,348 | 7,606 | 99.4 | 97.6 |
| Madagascar | 2008-09 | 93.6 | 8,569 | 12,448 | 98.8 | 95.6 |
| Malawi | 2015-16 | 63.5 | 13,448 | 17,286 | 99.2 | 97.7 |
| Mali | 2018 | 61.7 | 6,368 | 9,940 | 99.6 | 97.6 |
| Mozambique | 2011 | 73.1 | 7,623 | 11,102 | 99.8 | 99.1 |
| Myanmar | 2015-16 | 54.4 | 3,867 | 4,815 | 97.8 | 95.8 |
| Namibia | 2013 | 78.6 | 3,974 | 5,046 | 96.9 | 91.4 |
| Nepal | 2016 | 78.0 | 4,006 | 5,038 | 98.5 | 98.3 |
| Nigeria | 2018 | 52.8 | 21,792 | 33,924 | 99.4 | 99.3 |
| Pakistan | 2017-18 | 68.2 | 8,287 | 12,708 | 98.9 | 94.3 |
| Philippines | 2017 | 73.5 | 7,992 | 10,551 | 98.7 | 97.6 |
| Rwanda | 2014-15 | 80.5 | 5,955 | 7,856 | 99.9 | 99.5 |
| Senegal | 2010-11 | 74.5 | 8,147 | 12,330 | 98.4 | 92.7 |
| Sierra Leone | 2013 | 81.1 | 8,524 | 11,938 | 99.3 | 97.2 |
| South Africa | 2016 | 70.4 | 3,036 | 3,548 | 83.4 | 86.2 |
| Tajikistan | 2017 | 60.3 | 4,238 | 6,195 | 99.1 | 99.2 |
| Tanzania | 2015-16 | 44.9 | 7,050 | 10,233 | 98.4 | 97.3 |
| Timor-Leste | 2016 | 65.0 | 4,923 | 7,221 | 98.6 | 97.0 |
| Togo | 2013-14 | 81.4 | 5,016 | 6,979 | 99.1 | 97.8 |
| Uganda | 2016 | 58.3 | 10,263 | 15,522 | 98.2 | 97.0 |
| Zimbabwe | 2015 | 64.9 | 4,833 | 6,132 | 98.8 | 96.2 |

$\phi$ denotes the proportion of children having received a supplement of Vitamin A within the last 6 months (in %).

VAS status of the child was obtained from the mother via the woman’s survey; women between the ages of 15-49 were eligible to participate. Our data set included one record for every child of an interviewed woman, born in the five years preceding the survey. The outcome of interest was VAS coverage, i.e., *ϕ*, with *ϕ* being the proportion of children receiving a VAS dose in the six months preceding the survey within a primary sampling unit (PSU). We only included surveys which contained complete information about VAS for all children in the household.

For each cluster (a group of neighboring households based on geographic location), we extracted the outcome variable (i.e., VAS coverage) together with the location of the of the clusters. The outcome variable was adjusted by the individual weight for women and the household weight multiplied by the inverse of the individual response rate for women in the stratum. Strata are defined by geographic region and by urban/rural areas within each region. We took the average for each stratum to calculate the proportion of children who received a VAS dose (*ϕ*).

### 2.2 Spatial covariates

Table 2 presents covariates included in our analytical model. To inform our model, we compiled a set of covariates from the following domains, aggregated to a 5x5 *km*^2^ pixel level:

**Table 2:** Overview of variables

| Variable | Description |
| --- | --- |
| VIT.prop | $\phi$, with $\phi$ being the proportion of children having received a Vitamin A supplement within the last 6 months (year dependent on DHS survey) |
| ACCESS | Travel time to nearest city (2015) |
| ARIDITY | Average yearly precipitation divided by the average yearly potential evapotranspiration (2015) |
| POPULATION | 100m resolution population count (for all ages) using constrained top-down methods and adjusted to match United Nations national population counts (2020) |
| FROST | Average number of frost days (2015) |
| GROWPERIOD | Average number of days between greenup and dormancy (January 1982-December 2020) |
| LIVESTOCK | Aggregated count of livestock (2010) |
| MFAD | Modified Functional Attribute Diversity Index (using data from 1993-2003 for crop production and 2000-2005 for livestock production) |
| NONVEGETATED | Average proportion of areas not vegetated (January 2000 - December 2020) |
| SOILMOISTURE | Average soil moisture 10-40 cm underground from (January 1982 - December 2020) |
| SOILTEMPERATURE | Average soil temperature 10-40 cm underground (from January 1982 - December 2020) |
| B12YIELD | Production of vitamin B 12 through crop and livestock (using data from 1993-2003 for crop production and 2000-2005 for livestock production) |
| VITAMINAYIELD | Production of vitamin A through crop and livestock (using data from 1993-2003 for crop production and 2000-2005 for livestock production) |

Note: Removed covariates by country:
Egypt: FROST, Bangladesh: FROST, Haiti: SOILMOISTURE, Zimbabwe: ARIDITY, Cameroon: VITAMINAYIELD, WORLDPOP, Burundi: MFAD, NONVEG, Guinea: LIVESTOCK, MFAD, Liberia: LIVESTOCK, MFAD, Myanmar: FROST, VITAMINAYIELD, WORLDPOP, Ethiopia: VITAMINAYIELD, WORLDPOP

Geographic: we included travel time to the nearest city (Pfeffer et al 2018) and the total population count per pixel (Worldpop 2020b).

Climatic: we included average annual aridity and frost (Harris et al 2020), growing period (Friedl et al 2019), proportion of non-vegetated regions (DiMiceli et al 2015), and average soil temperature and soil moisture (NASA GSFC Hydrological Sciences Laboratory 2018). For the growing period and the non-vegetated regions, we calculated the average from January 2000 to December 2020.

Nutritional: we included the aggregated amount of livestock (Gilbert et al 2018a,b,c,d,e,f) and the Modified Functional Attribute Diversity Index (MFAD), which reflects the nutritional diversity produced in each 5x5 *km*^2^ pixel (Herrero et al 2017). Our analysis also accounted for the nutritional yield of vitamin B12 and A (Herrero et al 2017), as calculated through the average production of crops and livestock per grid cell.

For simplicity, we use the most recent values available for all covariates. All covariates and the outcome variable were scaled multiplicatively to values between 0.0001 to 0.9999, which prevents numerical instability during model fitting (Gómez Rubio 2020), allows for non-linear transformation of the covariates, and facilitates the comparison of estimates across covariates. Due to insufficient data quality in some cases, which often concurred with numeric instability, we omitted some covariates in selected countries.

### 2.3 Statistical analysis

We used a Bayesian hierarchical modeling framework to model VAS coverage across countries. We applied the following model:

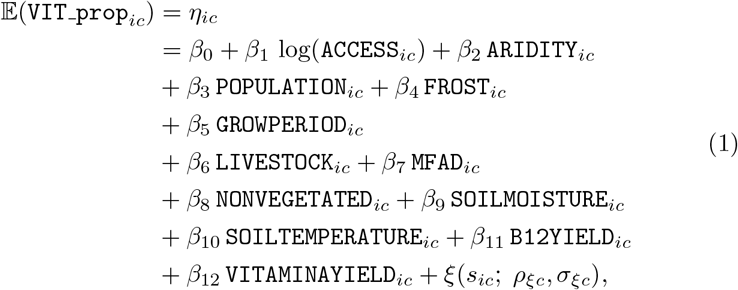

where 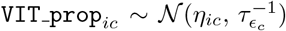 is *ϕ*, observed in PSU *i* in country *c*. This value describes VAS coverage. We assumed that `VIT_prop`_*ic*_ follows a Gaus-sian distribution with expectation *η*_*ic*_ and precision of the Gaussian errors *τ*_*ϵc*_.

While a binomial model seems an obvious choice, there is currently no methodological approach to take into account the weighting of PSUs; thus, we used the weighted proportion of VAS, as defined in *ϕ*.

Aside from the selected covariates which we assumed to be related to VAS coverage, our model used a spatial Gaussian process, *ξ*(*s*_*ic*_; *ρ*_*ξc*_, *σ*_*ξc*_) to capture spatial dependencies. We considered spatial autocorrelation explicitly by modelling the covariance structure of our residuals in space. Through this procedure, we captured the correlation structure of the data generating process to increase predictive accuracy of VAS coverage for locations where data is scarce. For this purpose, we applied a stationary Gaussian field with a Matérn covariance function:

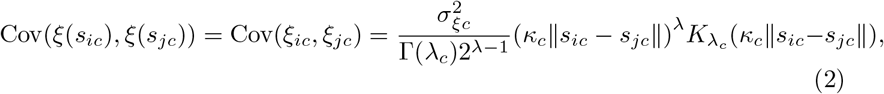

where 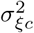 denotes the marginal variance of the Gaussian random field, ∥*s*_*ic*_ − *s*_*jc*_∥ is the Euclidean distance between two locations *s*_*ic*_, *s*_*jc*_ ∈ ℝ^2^, 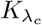 describes the modified Bessel function of second kind and order *λ*_*c*_ *>* 0 (accounting for the degree of smoothness of the spatial process), and *κ*_*c*_ *>* 0 is the scaling parameter associated with range *ρ*_*ξc*_ which can be interpreted as the distance above which we consider the spatial effect as negligible (Lindgren and Rue 2015).

The stochastic partial differential equation (SPDE) (Lindgren and Rue 2015) approach offers a computationally efficient implementation for our model. This approach applies Integrated Nested Laplace Approximations (INLA) (Rue et al 2009) and is easily applicable with the `R`-package `R-INLA`. This package has been very popular and successful, particularly in epidemiological modelling (Cameletti et al 2011, Bhatt et al 2015, Seufert et al 2022). `R-INLA` not only caters to a plethora of models (Rue et al 2009), but is also often faster than models using Markov Chain Monte Carlo, and provides easily interpretable diagnostics (Wang et al 2018).

Due to our lack of prior understanding about the extent and size of the spatial structure of VAS coverage in each country, we attempted to capture the spatial effects with penalized complexity priors, as suggested by Fuglstad et al (2017) and Simpson et al (2017). We imposed penalized complexity priors on both hyperparameters *ρ*_*ξc*_ and *σ*_*ξc*_ which are associated with the spatial random field *ξ*_*c*_. Therefore, we needed to define a threshold and a probability stemming from prior knowledge for which the true values of each considered parameter were above or below this threshold. We selected the following weakly informative priors on the range *ρ*_*ξc*_ and marginal variance 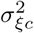 of *ξ*_*c*_: *P* (*ρ*_*ξc*_ *<* 0.01 *× δ*_*max*_) = 0.01, with *δ*_*max*_ describing the maximum Euclidean distance between two PSUs across the country and *P* (*σ*_*ξc*_ *>* 1) = 0.01. Further details on the prior specifications are provided in the Appendix.

Estimates of the prevalence of VAS coverage are presented as means with Bayesian uncertainty intervals (i.e., credible intervals) at a 5x5 *km*^2^ pixel level. We also present VAS coverage in terms of the absolute population size by multiplying the prevalence of VAS coverage with the total population of children under the age of five in each 5x5 *km*^2^ pixel (Worldpop 2020a). This data is useful for guiding planning and resource allocation. Lastly, we present exceedance probabilities, which indicate the probabilities of VAS coverage estimates being above or below a given threshold value. Exceedance probabilities are useful for assessing unusually low VAS coverage and also highlight areas where the number of observations is scarce. Thus, exceedance maps may be informative for policymakers to identify areas that have very low VAS coverage or where more data is required. The probability that the VAS coverage in pixel *i* in country *c* is higher than a value *h* can be expressed as *P* (*θ*_*ic*_ *> h*). This probability can be calculated by subtracting *P* (*θ*_*ic*_ ≤ *h*) from 1 as follows:

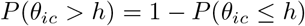

In grid cells with probabilities close to 1, it is highly likely that the VAS coverage estimate exceeds the threshold. Pixels with probabilities close to 0 depict regions where it is highly unlikely that the VAS coverage estimate exceeds the threshold. Grid cells with probabilities around 0.5 have the highest uncertainty and indicate regions where the coverage is below or above the threshold with equal probability. We chose two thresholds of *h* = 0.5 and *h* = 0.7; thus, our estimates indicate the probability that VAS coverage exceeds 50% or 70% respectively, for a pixel *i* in country *c*.

Validation of model fit and model specification were performed through comparison of the Watanabe–Akaike information criterion and through conditional predictive ordinates.

## 3 Results

The 45 countries included in our data set spanned the following UNICEF regions: Latin America and the Caribbean (LAC, *n* = 4), South Asia (*n* = 4), East Asia and the Pacific (*n* = 4), Central Asia (*n* = 2), Middle East and North Africa (*n* = 1), West and Central Africa (*n* = 15), and Eastern and Southern Africa (*n* = 15). Survey years ranged from 2006 - 2018. All countries had high response rates (see Table 1). For each continent, we present maps with the following results on a 5x5 *km*^2^ pixel level: mean estimates of VAS coverage, absolute VAS coverage (i.e., the mean estimates of VAS coverage multiplied by the population of children under the age of five (Worldpop 2020a)), the credible interval width of our estimates, and the exceedance probability for thresholds of *h* = 0.5 and *h* = 0.7.

### 3.1 Latin America and the Caribbean

Figures 1a and 1b show the estimated VAS coverage as a proportion and in absolute numbers, in the LAC countries examined. VAS coverage was higher in Honduras and Guatemala (∼ 50-60%) relative to Haiti and Bolivia (∼ 20-30%). In all four countries, there were slight sub-national variations in VAS coverage. In terms of absolute numbers, VAS coverage did not vary substantially either between or within countries. This is likely due to the demographic structure of the countries examined, characterized by a few densely populated metropolitan centers and many sparsely populated areas. The highest coverage in absolute terms was in Guatemala and the lowest coverage was in Bolivia.

**Fig. 1:**
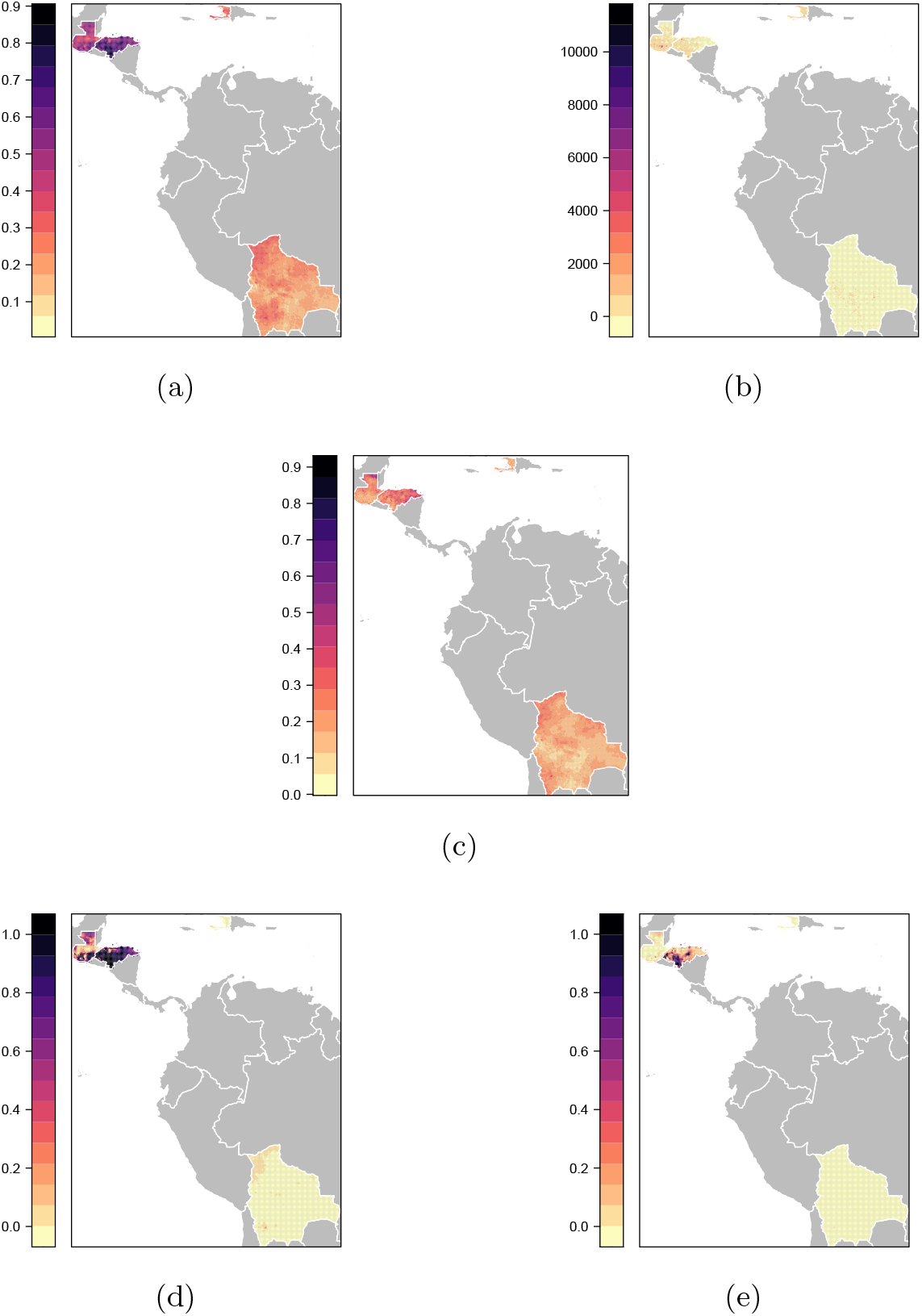
Latin America and the Caribbean: **(a)** Estimated mean **(b)** Estimated mean adjusted by population of children under 5 **(c)** 95% Credible Interval width **(d)** Exceedance probability h = 0.5 **(e)** Exceedance probability h = 0.7

Figure 1c shows the 95% credible interval width for our VAS coverage estimates for the LAC countries. Interval width was widest in Honduras and in some 5x5 *km*^2^ pixels in Guatemala, indicating a greater degree of uncertainty about our VAS coverage estimates in these areas; whereas, in Bolivia and Haiti, the intervals were relatively narrow, implying greater certainty about our estimates.

Exceedance probabilities are presented in Figures 1d and 1e. At *h* = 0.5, we are confident that our estimates lie above this threshold in Honduras and below this threshold in Haiti and Bolivia. In Guatemala, the degree of certainty regarding whether our estimates exceed this threshold varied substantially by region. For a *h* = 0.7, we are very certain that our estimates lie below this threshold in Bolivia, Haiti, Guatemala, and most of Honduras.

### 3.2 Africa

Figure 2a shows that in most countries we examined in Africa, estimated VAS coverage was below 70%. Additionally, there were marked sub-national variations in coverage. Madagascar and Namibia demonstrated the highest coverage (*>*70%), while Egypt and Chad had the lowest coverage (*<*40%). Coverage was higher and more uniform in southern African countries relative to countries in other regions of the continent. In terms of absolute population sizes, estimated VAS coverage was high in several west African countries (e.g., Nigeria, Benin, Togo, Burkina Faso, Ghana, Côte d’Ivoire). Absolute coverage was also relatively high in many parts of Ethiopia, Uganda, Malawi, and Madagascar in eastern and southern Africa (see Figure 2b).

**Fig. 2:**
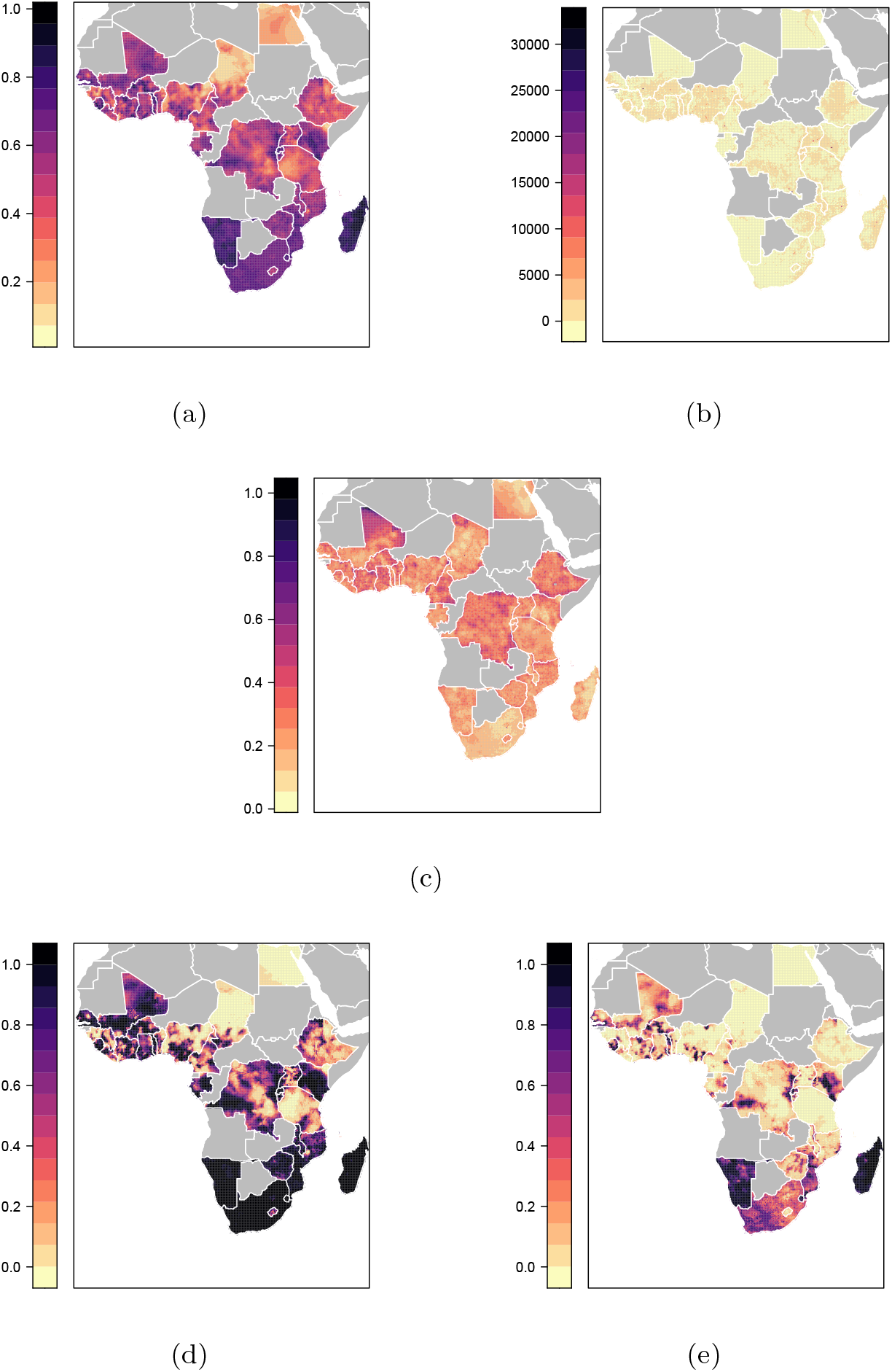
Africa: **(a)** Estimated mean **(b)** Estimated mean adjusted by population of children under 5 **(c)** 95% Credible Interval width **(d)** Exceedance probability h = 0.5 **(e)** Exceedance probability h = 0.7

As shown in Figure 2c, credible intervals were moderately wide for several countries (e.g., Mali, Cameroon, the DRC, and Ethiopia), indicating greater uncertainty about our VAS coverage estimates for these countries. Interval width was narrowest for South Africa and Madagascar, suggesting greater certainty about our estimates for these countries.

Figures 2d and 2e show exceedance probabilities for our estimates for African countries. At *h* = 0.5, there is a high degree of certainty that our estimates for VAS coverage exceed this threshold in southern African countries (i.e., South Africa, Namibia, Zimbabwe, Eswatini) and some east African countries (e.g., Madagascar, Mozambique, and Malawi). Additionally, the probability that our estimate exceeds this threshold was high in large parts of Kenya, and in sub-national regions of some countries (e.g., south-eastern Gabon, southern Nigeria). We can be certain that our estimates for Egypt and Chad fall below this threshold. However, in many countries, there were several areas where we are uncertain whether our estimates fall above or below the threshold (e.g., northern Mali, northern Ghana, south-eastern Ethiopia, northern DRC). At *h* = 0.7, we can be very certain that our estimates of VAS coverage lie above this threshold for Madagascar, Eswatini and Namibia, and below this threshold for most countries. In South Africa, Mozambique, Kenya, and Mali, we are most uncertain as to whether our estimates fall above or below 70%.

### 3.3 Asia

As shown in Figure 3a, most countries we examined had relatively moderate levels of estimated VAS coverage (*>*50%). Nepal and the Philippines had the highest coverage, and the Kyrgyz Republic had the lowest coverage. India and Pakistan had substantial sub-national variations in coverage. In India, northern and north-eastern regions had lower coverage relative to other parts of the country. In Pakistan, the south-western region had lower coverage relative to other regions. In terms of absolute numbers, as expected, estimated VAS coverage was highest in densely populated South Asian countries (e.g., India, Bangladesh) and the Philippines, with particular outliers in the urban agglomerations. Coverage was lowest in sparsely populated Central Asian countries (i.e., Tajikistan and the Kyrgyz Republic) (see Figure 3b).

**Fig. 3:**
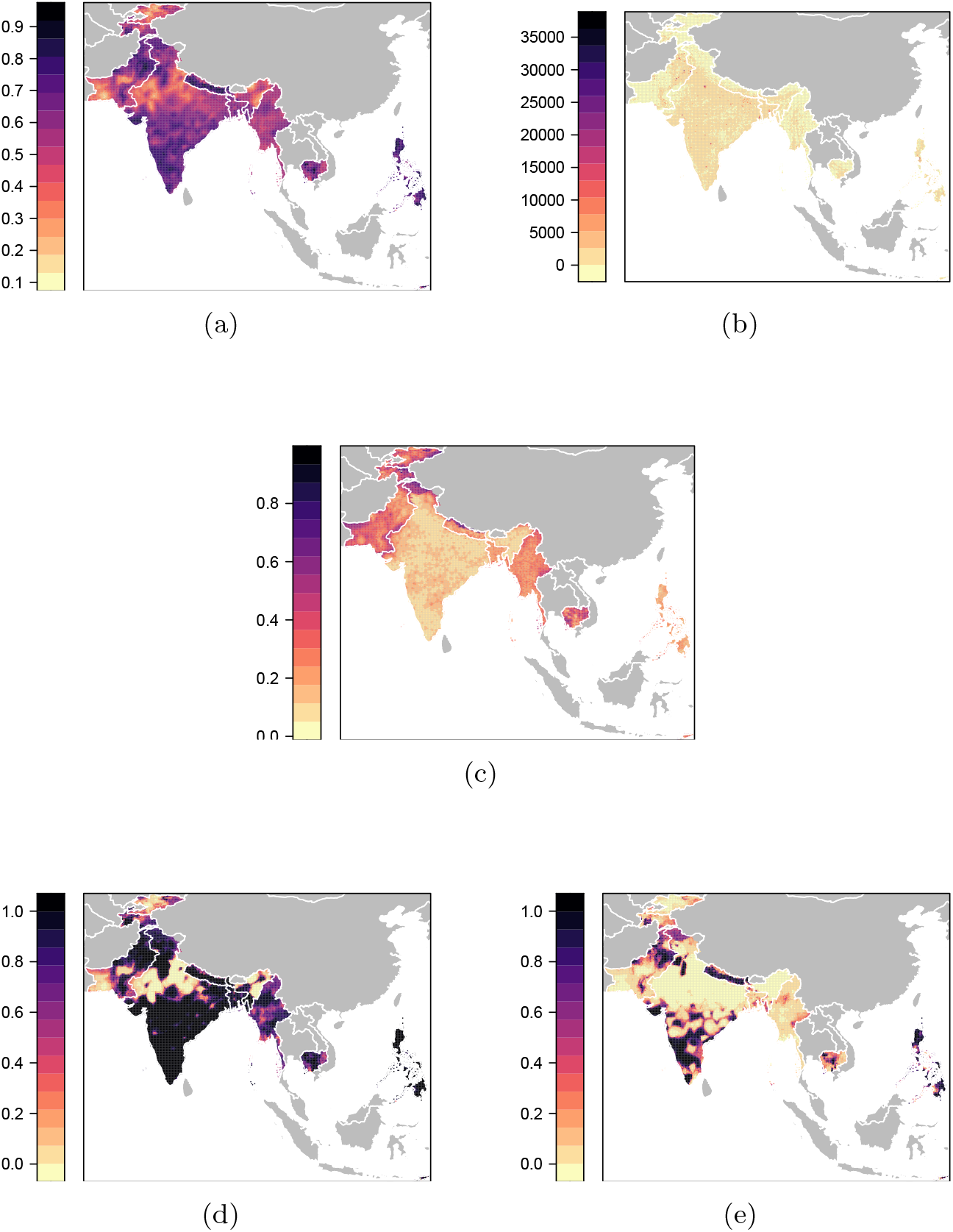
Asia: **(a)** Estimated mean **(b)** Estimated mean adjusted by population of children under 5 **(c)** 95% Credible Interval width **(d)** Exceedance probability h = 0.5 **(e)** Exceedance probability h = 0.7

Credible interval widths for our estimates are shown in Figure 3c. Wider interval widths for the Kyrgyz Republic, Tajikistan, Pakistan, Myanmar, and Cambodia indicate a higher degree of uncertainty about our estimates for these countries; whereas, credible interval width was generally narrow across India with the exception of Jammu and Kashmir, indicating greater certainty about our VAS coverage estimates across India.

Figure 3d shows that there is a high degree of certainty that our estimates for VAS coverage exceed 50% in the Philippines, Nepal, Bangladesh, most of India, and north-eastern Pakistan. In northern and northeastern India, we are certain that our estimates fall below 50%. In south-western Pakistan and in several areas of Cambodia, Myanmar, Tajikistan, and the Kyrgyz Republic, we are uncertain whether our estimates are above or below 50%. As shown in Figure 3e, in Nepal, southern India, northern Pakistan and parts of the Philippines, we are certain that our estimates exceed 70%; whereas, in northern and north-eastern India, south-western Pakistan, and the Kyrgyz Republic, we are certain that our estimates do not exceed 70%. In Cambodia, we are most uncertain about whether our estimates for VAS coverage fall above or below the threshold of *h* = 0.7.

## 4 Discussion

We examined VAS coverage at a fine geographic level (5x5 *km*^2^ pixels) between 2006–2018 in 45 UNICEF designated VAS priority countries to identify national and sub-national disparities in coverage. Of the countries examined, we found that the vast majority had coverage levels below 70%. Additionally, VAS coverage varied between and within countries; inter-national variations in coverage were most notable in LAC and Africa, and intra-national variations were greatest in Pakistan, India, and several western and central African countries. These findings underscore the need for all countries to scale up VAS coverage. Additionally, areas with especially low coverage must be prioritized to ensure equity.

Disparities in VAS coverage between countries could be attributable to differences in VAS distribution mechanisms. Many countries were initially able to achieve high levels of VAS coverage by integrating VAS into polio supplementary immunization activities (SIAs) (Chehab et al 2016, UNICEF 2018). However, with progress in polio eradication, countries that primarily relied on polio SIAs lost their main VAS delivery platform. We found higher levels of VAS coverage in southern and eastern African countries, which were less reliant on polio SIAs, whereas western and central African countries, which mainly relied on polio SIAs, had lower VAS coverage (UNICEF 2018). By developing flexible distribution mechanisms, countries can minimize drops in coverage when one distribution mechanism is affected (Chehab et al 2016, UNICEF 2018).

Political, sociocultural, and environmental factors may also contribute to disparities in VAS coverage, particularly at the sub-national level (Haselow et al 2022, Zegeye et al 2022). For example, our maps point to low VAS coverage in several conflict-affected areas, such as northeastern Nigeria and southwestern Pakistan. Consistent with a previous analysis, we also found low VAS coverage in Manipur and Nagaland states in north-eastern India, which have high poverty levels and are geographically remote (Bora 2022). Nutrition and immunization programs that have been successfully implemented in low-resource and complex settings offer insight into enabling factors. These include strong political commitment and leadership, microplanning at the district or administrative level, sustainable supply of VAS, flexible delivery mechanisms, social mobilization and communication, and training, supervision, and monitoring (Haselow et al 2022, Houston 2003, Rah et al 2014). Additionally, strategies for successfully implementing programs in conflict settings include negotiating secure physical access, engaging local communities, coordinating with other health and humanitarian aid delivery, delivering services at refugee camps and other mass gathering sites, collaborating with military and security personnel, and mobilizing during tranquil periods (Nnadi et al 2017).

Our high-resolution maps present a granular picture of the state of VAS coverage globally and can guide efforts to reduce national and sub-national disparities in coverage. As VAS need may vary across sub-national regions, these maps should be used in conjunction with granular data on VAD (if available) or U5MR to target VAS programs even more precisely. This will ensure that VAS programs reach children with the greatest need. For instance, in analyses examining U5MR in India and Africa at the 5x5 *km*^2^ pixel level, grid cells with the highest U5MR closely corresponded with grid cells having the lowest VAS coverage in our analysis (e.g., northern and north-eastern India; north-eastern Nigeria, southern DRC) (Golding et al 2017, Dandona et al 2020). This suggests that targeting VAS efforts in these areas could have a large impact on child health and survival.

Strengths of this study are the use of nationally representative data and high response rates, which ensure that findings are generalizable within each country. Additionally, as DHS procedures and methodologies are standardized, findings are comparable across countries. However, this study also had some limitations that should be noted. VAS coverage estimates in some grid cells lacked precision due to insufficient sample sizes and should be interpreted with caution. Our maps providing credible interval widths and exceedance probabilities quantify this uncertainty. These maps can aid in interpretation of results and highlight areas where more data is needed. Although areas with the lowest VAS coverage are likely to have the highest need for VAS, this may not always be the case. To assess equity in coverage, these findings should be interpreted in conjunction with data on indicators of VAS need (e.g., VAD prevalence, U5MR). Data used for this analysis spanned a 12-year period and VAS coverage may have changed substantially in countries for which we relied on older data. DHS assesses VAS coverage based on self-report of receipt (by the caregiver) in the preceding six months and therefore does not reflect full (i.e., two-dose) coverage for a child. It is also possible that respondents could have confused VAS with other nutrition or immunization interventions (e.g., polio) (UNICEF 2018). Data quality for covariates varied by country, causing us to drop some covariates for some countries from our analysis. From a statistical standpoint, it would be desirable to model the outcome variable through a binomial likelihood while also taking into account the weighting of observations; however, as this approach was not methodologically feasible, we used an alternative approach.

VAS is a low-cost, lifesaving intervention, yet it is estimated that more than a third of children in need of VAS are not receiving it (UNICEF 2018). Our high-resolution maps provide a comprehensive picture of the state of VAS coverage across priority countries and highlight national and sub-national disparities in coverage. These data, especially when calibrated against high-resolution data on indicators of VAS need, could help VAS programs achieve equity.

## Data Availability

Data availability
Data from the DHS Program:see link
Geospatial data for estimating maps was obtained from
Travel time: see link
Population: see link
Livestock:see link
MFAD, Vitamin A, Vitamin B12: upon request from Herrero M, Thornton PK, Power B, et al (2017) Farming and the geography of
nutrient production for human use: a transdisciplinary analysis. The Lancet
Planetary Health 1(1):
Soil moisture, soil temperature: see link
Non-Vegetation percentage: see link
Vegetation period:see link
Aridity and frost: see link

https://dhsprogram.com/data/available-datasets.cfm

https://malariajournal.biomedcentral.com/articles/10.1186/s12936-018-2500-5

https://hub.worldpop.org/geodata/listing?id=75

https://dataverse.harvard.edu/

https://disc.gsfc.nasa.gov/datasets/FLDAS_NOAH01_C_GL_M_001/summary

https://lpdaac.usgs.gov/products/mod44bv006/

https://lpdaac.usgs.gov/products/mcd12q2v006/

https://crudata.uea.ac.uk/cru/data/hrg/

## Acknowledgments

Acknowledgments are not compulsory. Where included they should be brief. Grant or contribution numbers may be acknowledged.

Please refer to Journal-level guidance for any specific requirements.

## Declarations

- Funding
- Conflict of interest/Competing interests (check journal-specific guidelines for which heading to use)
- Ethics approval
- Consent to participate
- Consent for publication
- Availability of data and materials
- Code supporting the findings of this study are available within the paper and its Supplementary Information. Data is publicly available (see citations in paper).
- Authors’ contributions

## A Tables

## C Additional statistical information

We apply the standard flat priors suggested for the hyperparameters and the intercept which `R` − `INLA` applies by default. We use normally distributed priors (𝒩 (mean, variance)) for the remaining fixed effects as follows:

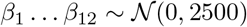

Furthermore, we use multivariate normally distributed priors on the spatial term *ξ*(_*ic*_):

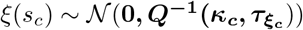

with precision matrix ***Q*** composed of the scaling parameter *κ*_*c*_ and GMRF precision 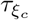. The precision matrix is specified in the Matérn covariance function. Without specific a priori knowledge on the unstructured errors, we use `R-INLA` default priors which assume log(*σ*_*ϵc*_^−1^) ∼ *logGamma*(1, 0.8).

## D Additional Tables

